# Age-dependent effects in the transmission and control of COVID-19 epidemics

**DOI:** 10.1101/2020.03.24.20043018

**Authors:** Nicholas G. Davies, Petra Klepac, Yang Liu, Kiesha Prem, Mark Jit, CMMID COVID-19 working group, Rosalind M Eggo

**Author notes:** these authors contributed equally (Petra Klepac, Yang Liu). correspondence to Rosalind M Eggo or Nicholas G Davies. Carl A B Pearson, Billy J Quilty, Adam J Kucharski, Hamish Gibbs, Samuel Clifford, Amy Gimma, Kevin van Zandvoort, James D Munday, Charlie Diamond, W John Edmunds, Rein MGJ Houben, Joel Hellewell, Timothy W Russell, Sam Abbott, Sebastian Funk, Nikos I Bosse, Fiona Sun, Stefan Flasche, Alicia Rosello & Christopher I Jarvis. Order of working group determined at random.

## Abstract

The COVID-19 pandemic has shown a markedly low proportion of cases among children. Age disparities in observed cases could be explained by children having lower susceptibility to infection, lower propensity to show clinical symptoms, or both. We evaluate these possibilities by fitting an age-structured mathematical model to epidemic data from six countries. We estimate that clinical symptoms occur in 25% (95% CrI: 19-32%) of infections in 10-19-year-olds, rising to 76% (68-82%) in over-70s, and that susceptibility to infection in under-20s is approximately half that of older adults. Accordingly, we find that interventions aimed at children may have a relatively small impact on total cases, particularly if the transmissibility of subclinical infections is low. The age-specific clinical fraction and susceptibility we have estimated has implications for the expected global burden of COVID-19 because of demographic differences across settings: in younger populations, the expected clinical attack rate would be lower, although it is likely that comorbidities in low-income countries will affect disease severity. Without effective control measures, regions with older populations may see disproportionally more clinical cases, particularly in the later stages of the pandemic.

---

**Extended Data Table 1.** Posterior estimates for the consensus susceptibility and clinical fraction from 6 countries. Note that susceptibility is a relative measure.

| *Parameter* | *Age Group* | *Mean* | *Quantile 2.5%* | *Quantile 25%* | *Quantile 50%* | *Quantile 75%* | *Quantile 97.5%* |
| --- | --- | --- | --- | --- | --- | --- | --- |
| *Susceptibility* | 0-9 | 0.33 | 0.25 | 0.3 | 0.33 | 0.36 | 0.43 |
| *Susceptibility* | 10-19 | 0.37 | 0.28 | 0.33 | 0.37 | 0.4 | 0.47 |
| *Susceptibility* | 20-29 | 0.69 | 0.54 | 0.64 | 0.69 | 0.74 | 0.82 |
| *Susceptibility* | 30-39 | 0.81 | 0.65 | 0.76 | 0.81 | 0.86 | 0.95 |
| *Susceptibility* | 40-49 | 0.74 | 0.59 | 0.7 | 0.75 | 0.79 | 0.86 |
| *Susceptibility* | 50-59 | 0.8 | 0.65 | 0.76 | 0.81 | 0.85 | 0.93 |
| *Susceptibility* | 60-69 | 0.89 | 0.72 | 0.85 | 0.9 | 0.94 | 0.99 |
| *Susceptibility* | 70+ | 0.77 | 0.62 | 0.72 | 0.78 | 0.82 | 0.9 |
| *Clinical fraction* | 0-9 | 0.4 | 0.31 | 0.36 | 0.4 | 0.44 | 0.48 |
| *Clinical fraction* | 10-19 | 0.25 | 0.19 | 0.23 | 0.25 | 0.29 | 0.32 |
| *Clinical fraction* | 20-29 | 0.37 | 0.28 | 0.33 | 0.37 | 0.41 | 0.46 |
| *Clinical fraction* | 30-39 | 0.42 | 0.33 | 0.38 | 0.42 | 0.46 | 0.51 |
| *Clinical fraction* | 40-49 | 0.51 | 0.41 | 0.47 | 0.51 | 0.55 | 0.6 |
| *Clinical fraction* | 50-59 | 0.59 | 0.48 | 0.55 | 0.59 | 0.62 | 0.67 |
| *Clinical fraction* | 60-69 | 0.72 | 0.63 | 0.69 | 0.72 | 0.75 | 0.79 |
| *Clinical fraction* | 70+ | 0.76 | 0.68 | 0.73 | 0.76 | 0.79 | 0.82 |

**Extended Data Table 2.** Model parameters.

| Parameter | Description | Applies in fits | Value | Reference |
| --- | --- | --- | --- | --- |
| $d_E$ | Incubation period (E to $I_P$ and E to $I_S$; days) | All | $\sim \text{gamma}(\mu = 3.0, k = 4)$ | Derived from<sup>1</sup><sub>2</sub> |
| $d_P$ | Duration of preclinical infectiousness (days) | All | $\sim \text{gamma}(\mu = 2.1, k = 4)$ | Derived from<sup>2</sup> |
| $d_C$ | Duration of clinical infectiousness ($I_C$ to R; days) | All | $\sim \text{gamma}(\mu = 2.9, k = 4)$ | <sup>3</sup> |
| $d_S$ | Duration of subclinical infectiousness (days) | All | $\sim \text{gamma}(\mu = 5, k = 4)$ | Assumed |
| $u_i$ | Susceptibility for age group $i$ | Varies by age in Wuhan hypothesis 2, otherwise all ages equal | Estimated | |
| $y_i$ | Probability of clinical infection for age group $i$ | Varies by age in Wuhan hypothesis 3, otherwise all ages equal | Either fixed (50%) or estimated | <sup>4</sup> |
| $f$ | Relative infectiousness of subclinical cases | All | 50% (0% and 100% in sensitivity analysis) | Assumed |
| $c_{ij}$ | Number of age-$j$ individuals contacted by an age-$i$ individual per day | All | Country-specific contact matrix (sensitivity analysis using synthetic matrices<sup>19</sup>) | China<sup>32</sup>; UK<sup>7</sup>; Zimbabwe<sup>34</sup> |
| $N_i$ | Number of age-$i$ individuals | All | Demographic data | <sup>5</sup> |
| $\Delta t$ | Time step for discrete-time simulation | All | 0.25 days | |
| $A_{min}, A_{max}$ | Age range of seed cases | Wuhan | Estimated | |
| $t_{seed}$ | Day upon which seeding of infections starts | All | Estimated | |
| $q_H$ | Relative change in non-school contacts during lunar new year holidays | Wuhan | Estimated | |
| $q_L$ | Relative change in non-school contacts following large-scale restrictions | Wuhan, South Korea, Shanghai, Beijing, Italy | Estimated | |
| $t_L$ | Day upon which large-scale restrictions start | Wuhan, South Korea, Shanghai, Beijing, Italy | Fixed to January 23 for Wuhan; estimated for other settings | |

**Extended Data Figure 1.**
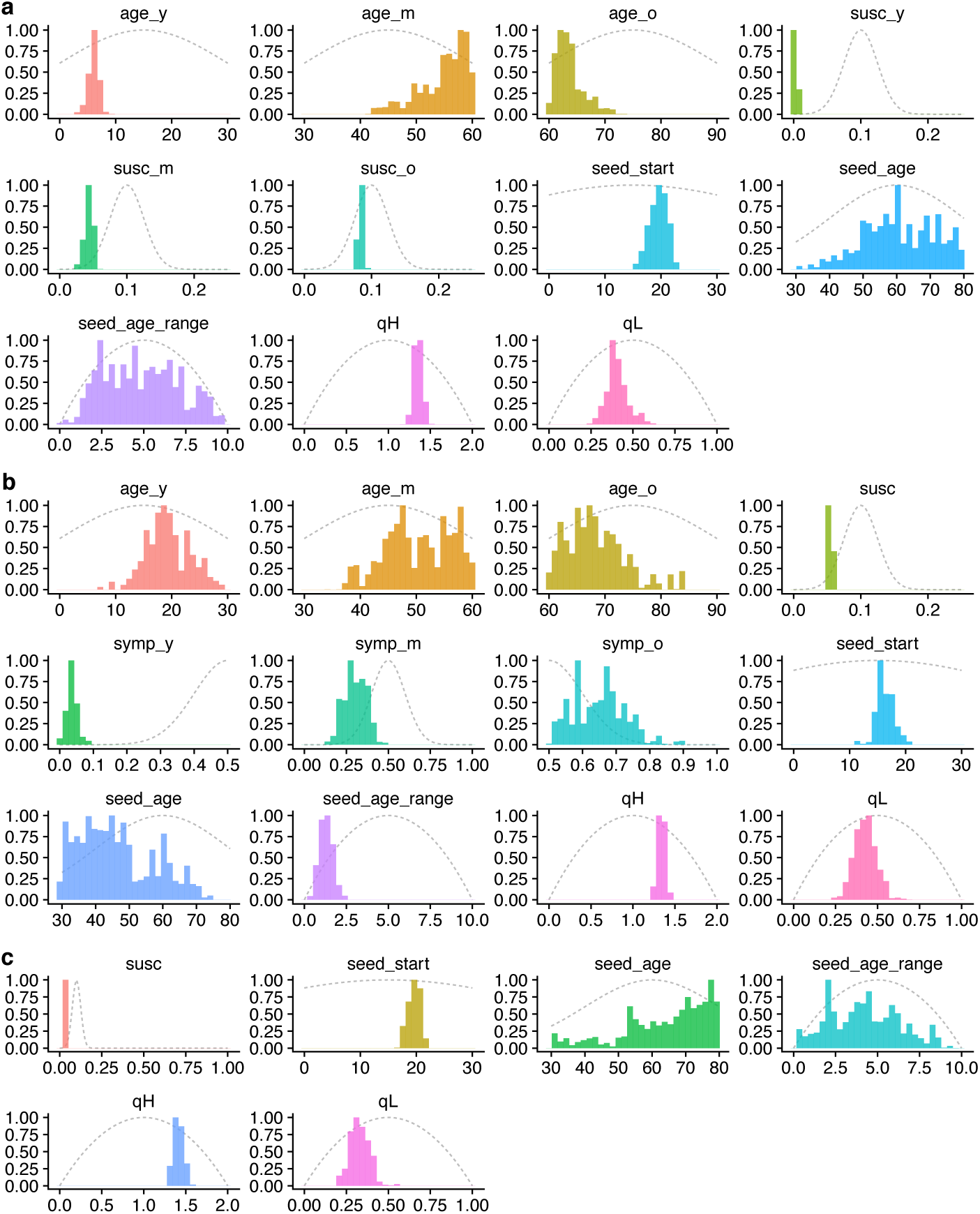
Prior distributions (grey dotted lines) and posterior distributions (coloured histograms) for model parameters fitting to the early epidemic in Wuhan (Fig. 1, main text); seed_start is measured in days after November 1st, 2019. **(a)** Model 1 (age-varying contact patterns and susceptibility); **(b)** Model 2 (age-varying contact patterns and clinical fraction); **(c)** Model 3 (age-varying contact patterns only). See also Supplementary Table 3.

**Extended Data Figure 2.**
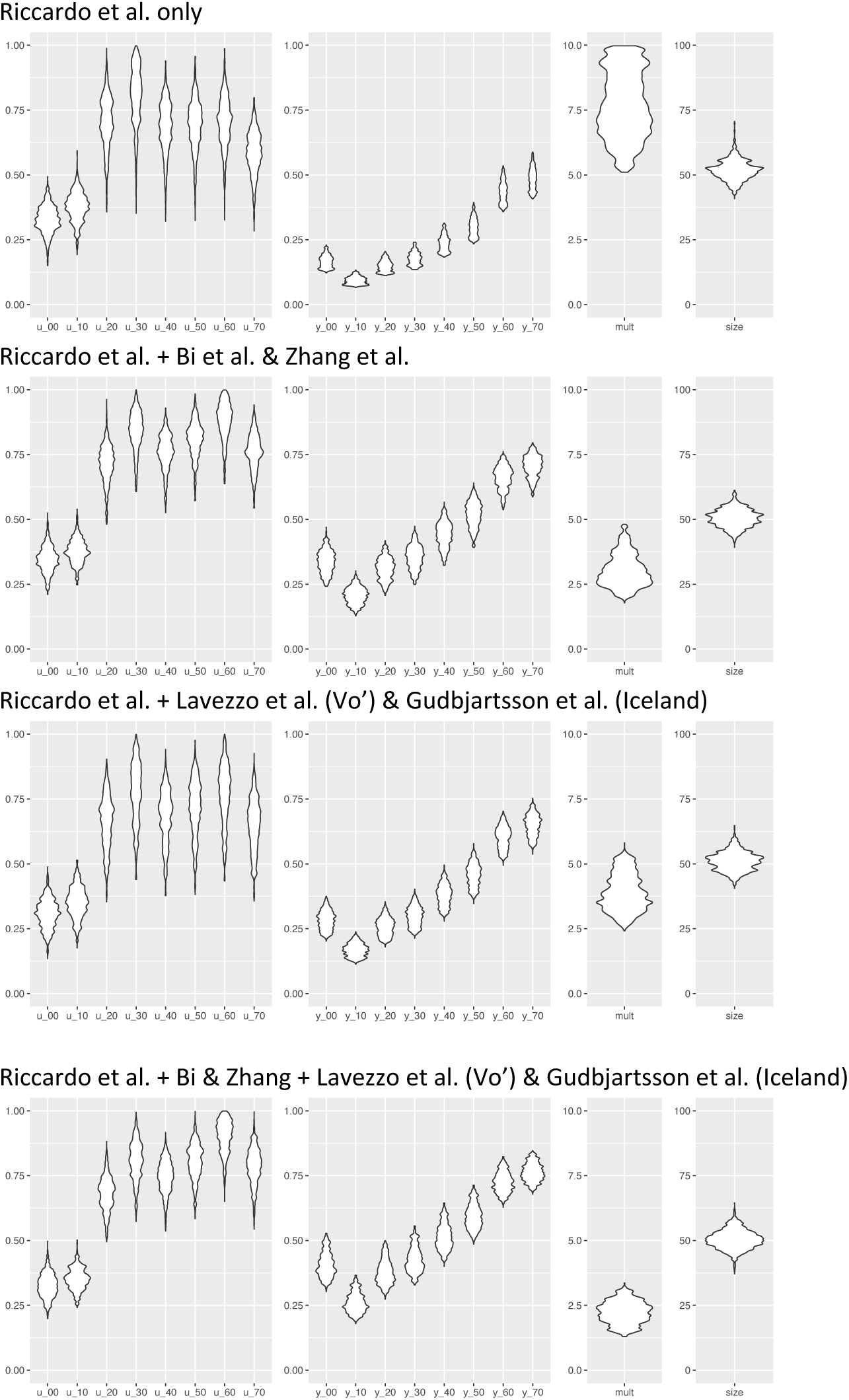
Analysis showing how the inferred age-varying susceptibility (first column) and age-varying clinical fraction (second column) depend upon the additional data sources used.

**Extended Data Figure 3.**
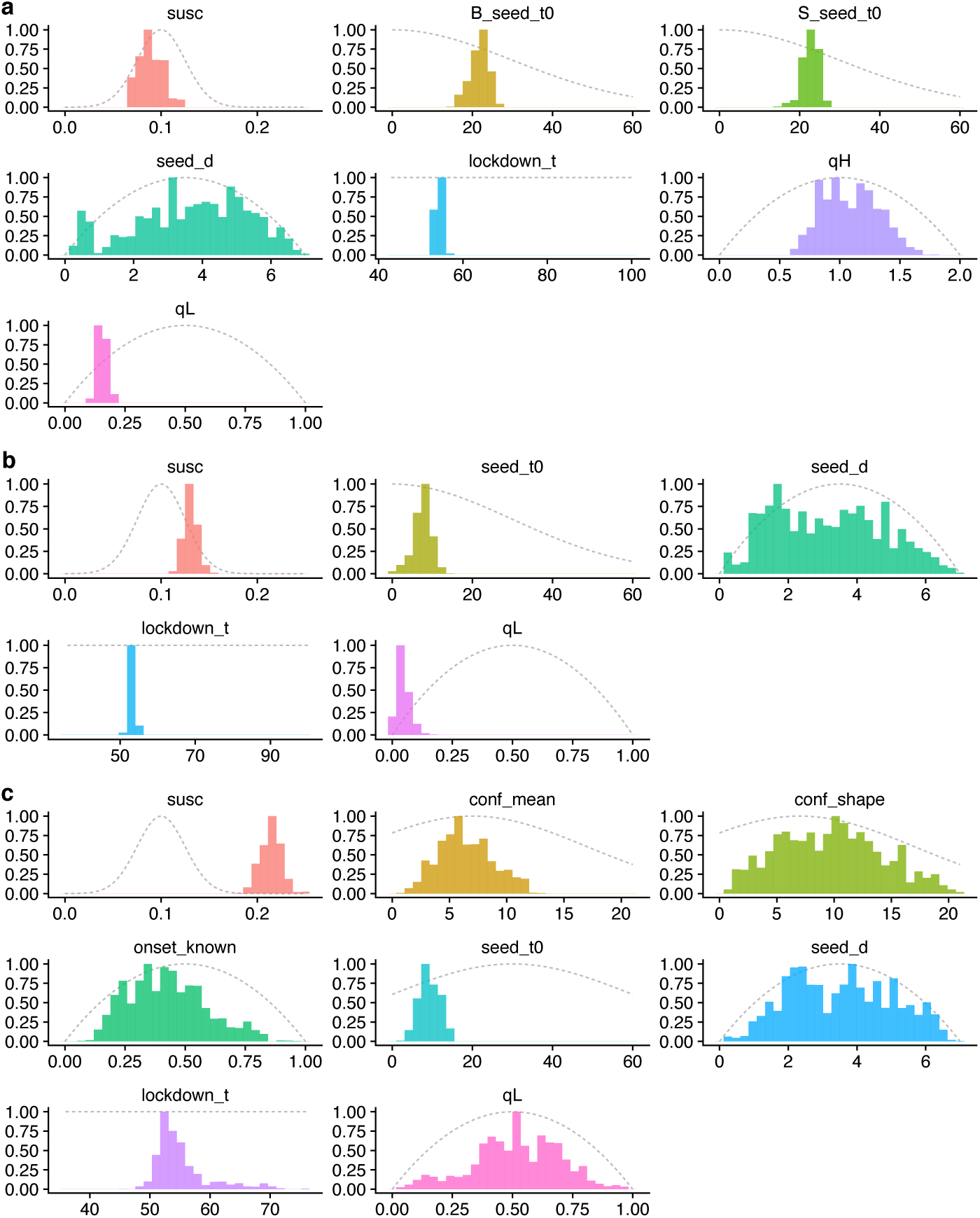
Prior and posterior distributions for the epidemics in **(a)** Beijing and Shanghai, **(b)** South Korea and **(c)** Lombardy using the “consensus” fit for age-specific clinical fraction and assuming subclinical infections are 50% as infectious as clinical infections (see Fig. 2c, main text). For **(a)**, times are in days after December 1st, 2019; for **(b)** and **(c)**, times are in days after January 1st, 2019. Note, seed_d is the inferred duration of the seeding event. See also Supplementary Table 3.

**Extended Data Figure 4.**
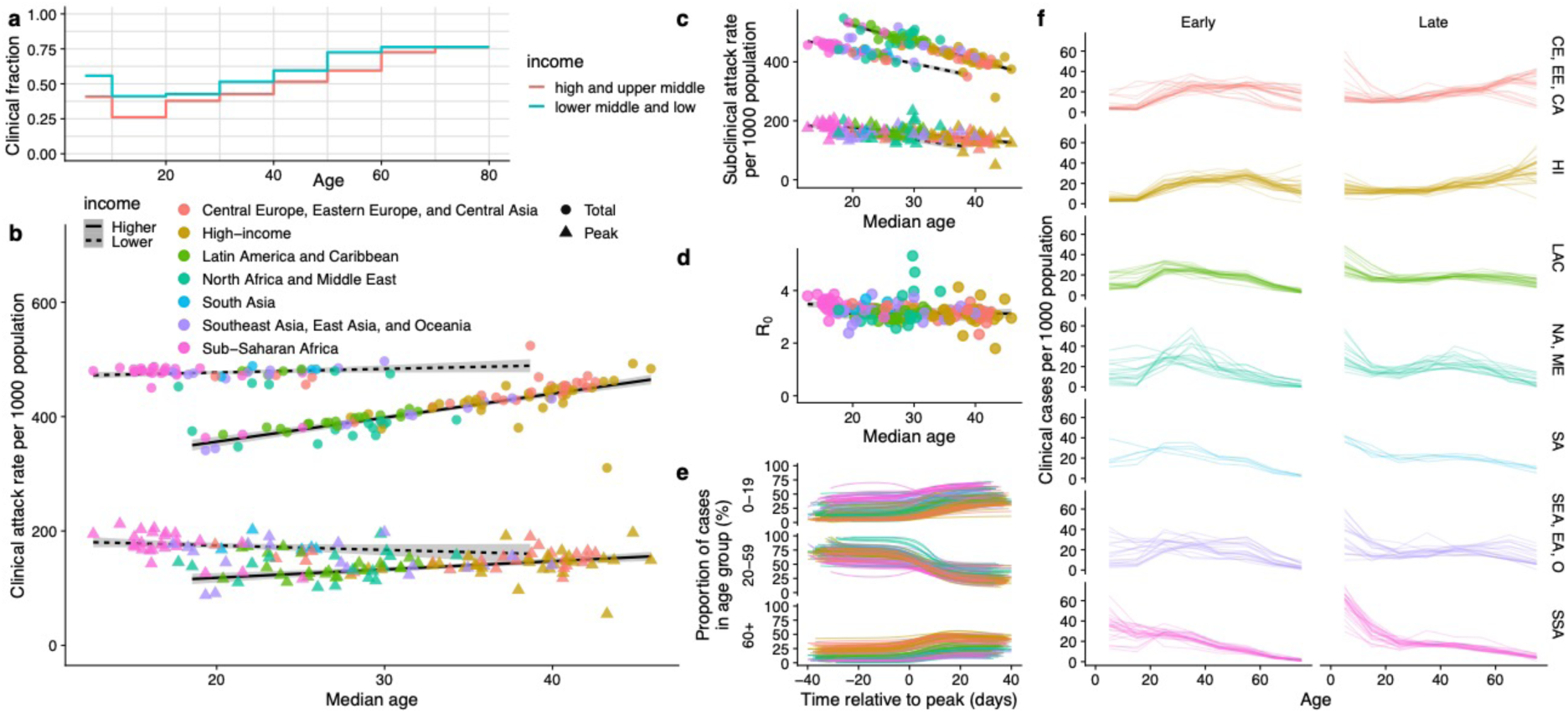
Global projections assuming greater severity in lower-income countries. **(a)** Schematic age-specific clinical fraction for higher-income and lower-income countries. **(b-f)** Illustrative results of the projections for 146 capital cities assuming a higher age-varying clinical fraction in lower-income countries. See Fig. 4 (main text) for details.

**Extended Data Figure 5.**
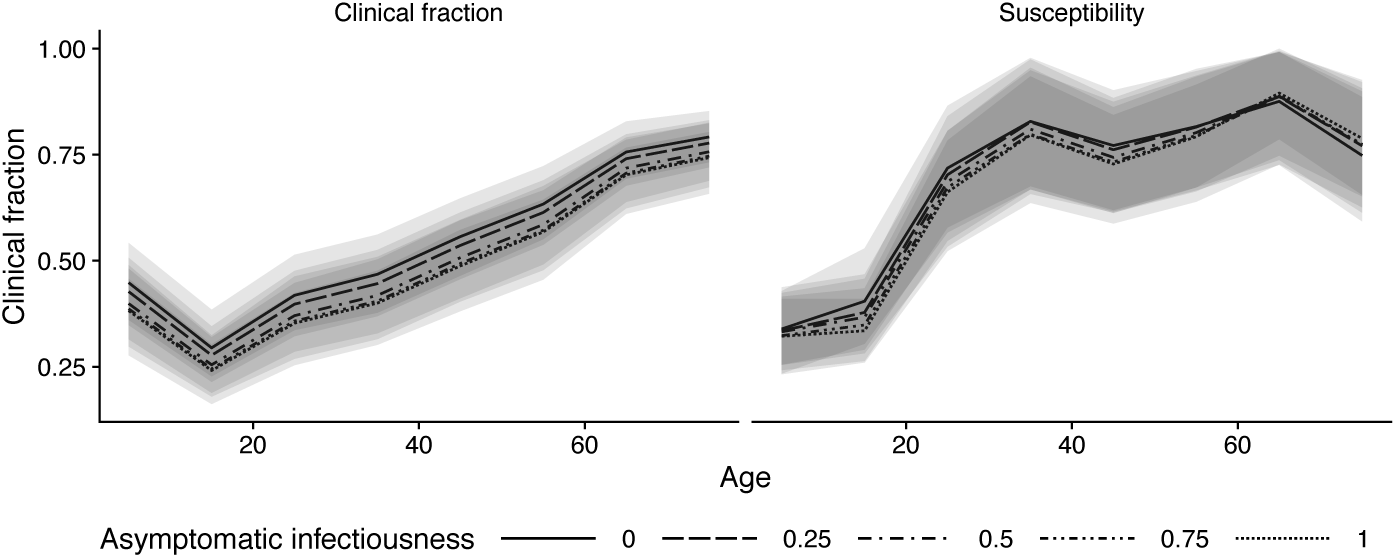
Consensus age-specific clinical fraction and susceptibility, assuming subclinical infections are 0%, 25%, 50%, 75%, or 100% as infectious as clinical infections.

**Extended Data Figure 6.**
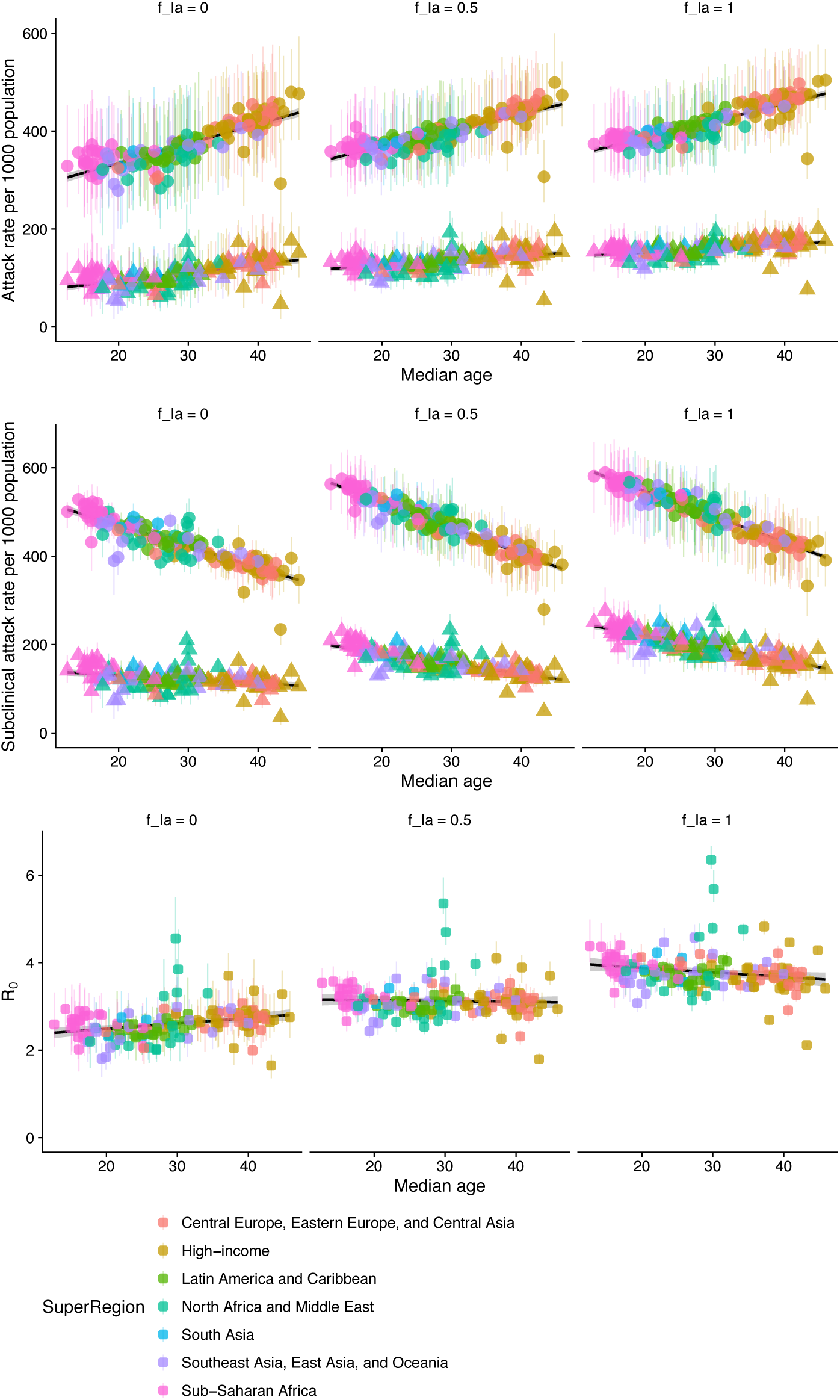
**(a)** Projected total and peak clinical case attack rate for 146 capital cities, under different assumptions for the infectiousness of subclinical infections. **(b)** Projected total and peak subclinical infection attack rate for 146 capital cities, under different assumptions for the infectiousness of subclinical infections. **(c)** Projected differences in R0 among 146 capital cities, under different assumptions for the infectiousness of subclinical infections.

**Extended Data Figure 7.**
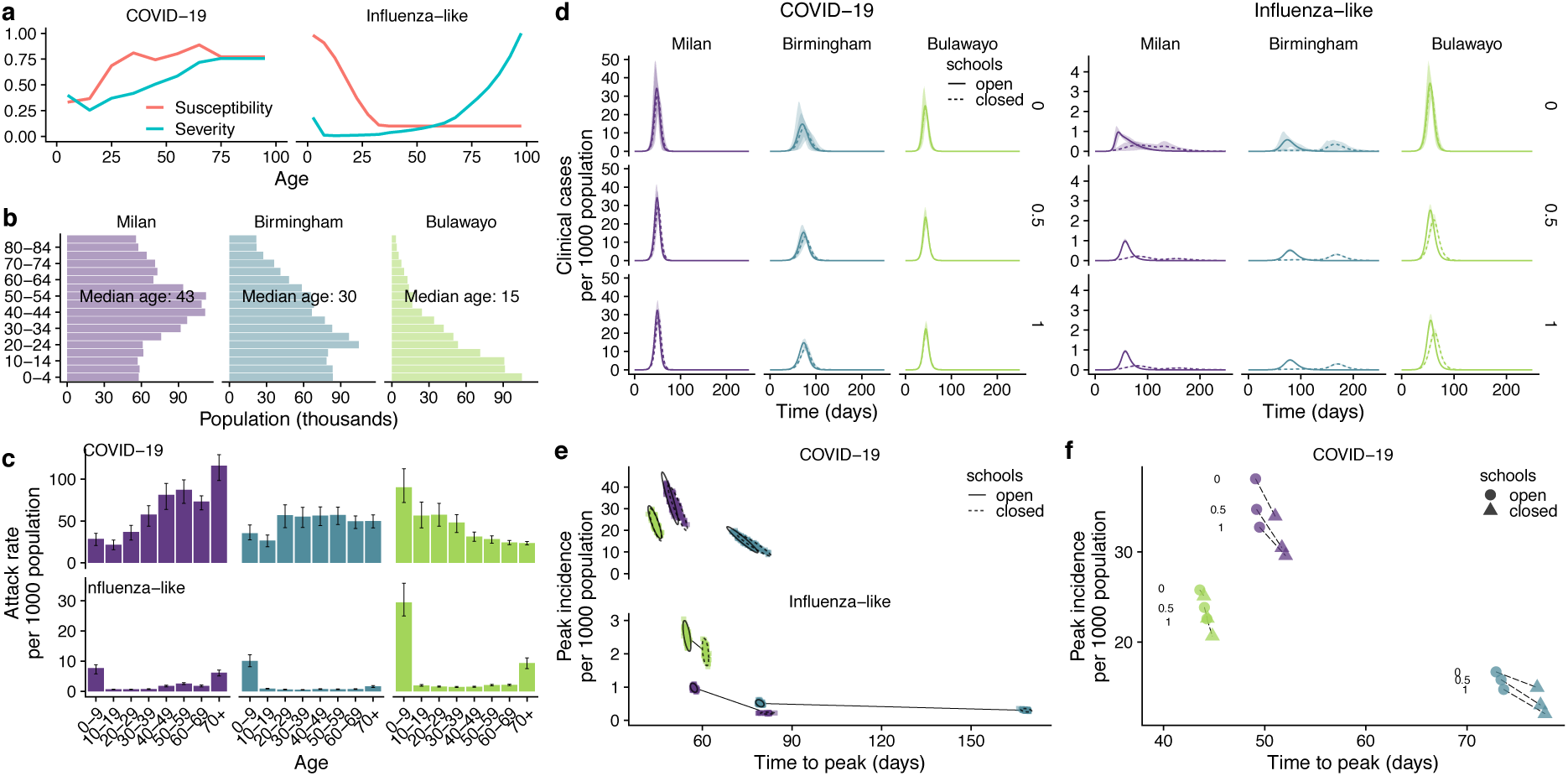
Comparison of school closures in three exemplar cities when susceptibility *u_i_* is fixed across settings instead of R0.

**Extended Data Figure 8.**
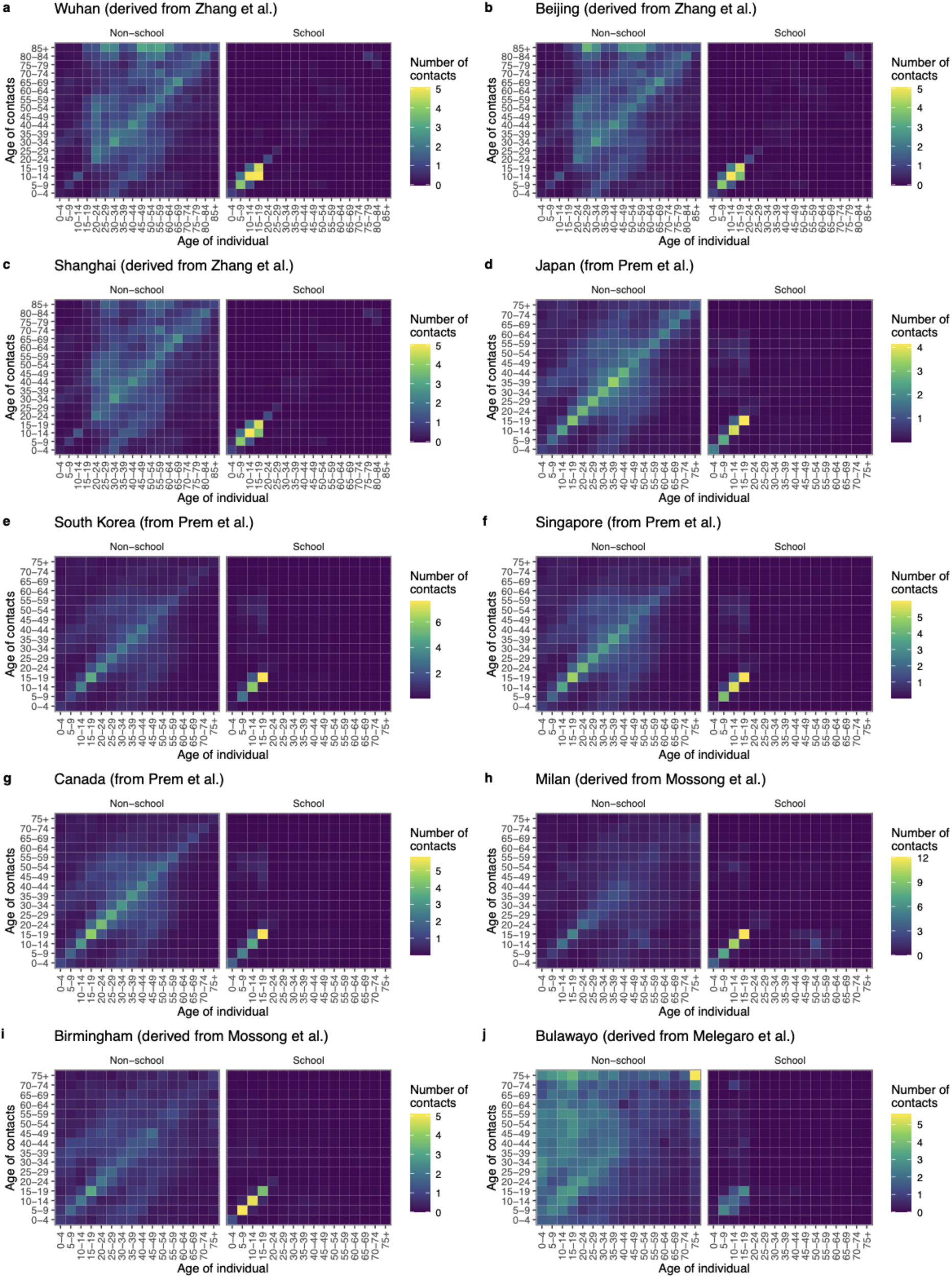
Contact matrices used for Figs. 1-3 of the main text. We have not shown matrices for all 12 regions of Italy modelled, nor for all 13 provinces of China modelled, as these show similar patterns to the matrices for Milan and for Wuhan, Beijing and Shanghai, respectively.

## Supplementary Information

**Supplementary Table 1.** Details of model fitting.

| Parameter | Description | Prior |
| --- | --- | --- |
| $u_i$ | Susceptibility to infection upon contact with an infectious person | Non-age-varying: $u_i \sim \text{normal}(\mu = 0.1, \sigma = 0.025, \min = 0)$<br>Age-varying: young, middle, and old age fit as<br>$a_y \sim \text{normal}(\mu = 15, \sigma = 15, \min = 0, \max = 30)$<br>$a_m \sim \text{normal}(\mu = 45, \sigma = 15, \min = 30, \max = 60)$<br>$a_o \sim \text{normal}(\mu = 75, \sigma = 15, \min = 60, \max = 90)$<br>Susceptibility for young, middle, and old age fit as<br>$u_y \sim \text{normal}(\mu = 0.1, \sigma = 0.025, \min = 0)$<br>$u_m \sim \text{normal}(\mu = 0.1, \sigma = 0.025, \min = 0)$<br>$u_o \sim \text{normal}(\mu = 0.1, \sigma = 0.025, \min = 0)$<br>Then<br>$u_i = \text{coss}(i\vert a_y, b_y, a_m, b_m, a_o, b_o)$ (see final row) |
| $y_i$ | Clinical fraction on infection | Non-age-varying: $y_i = 0.5$<br>Age-varying: young, middle, and old age fit as<br>$a_y \sim \text{normal}(\mu = 15, \sigma = 15, \min = 0, \max = 30)$<br>$a_m \sim \text{normal}(\mu = 45, \sigma = 15, \min = 30, \max = 60)$<br>$a_o \sim \text{normal}(\mu = 75, \sigma = 15, \min = 60, \max = 90)$<br>Susceptibility for young, middle, and old age fit as<br>$y_y \sim \text{normal}(\mu = 0.5, \sigma = 0.1, \min = 0, \max = 0.5)$<br>$y_m = 0.5$<br>$y_o \sim \text{normal}(\mu = 0.5, \sigma = 0.1, \min = 0.5, \max = 1)$<br>Then<br>$y_i = \text{coss}(i\vert a_y, y_y, a_m, y_m, a_o, y_o)$ (see below) |
| $t_{\text{seed}}$ | Timing of introduction of cases | $t_{\text{seed}} \sim \text{normal}(\mu = 15, \sigma = 30, \min = 0, \max = 30)$ |
| $q_H$ | Multiplicative factor for transmission during holiday period | $q_H \sim \text{beta}(\alpha = 2, \beta = 2)$ scaled to 0 – 2 |
| $q_L$ | Multiplicative factor for transmission during large-scale restrictions | $q_L \sim \text{beta}(\alpha = 2, \beta = 2)$ |
| $A_{\min}, A_{\max}$ | Age bounds for introduced cases | $A \sim \text{normal}(\mu = 60, \sigma = 20, \min = 40, \max = 80)$<br>$A_{\text{range}} \sim \text{beta}(\alpha = 2, \beta = 2)$ scaled to 0 – 10<br>$A_{\min} = A - A_{\text{range}}$<br>$A_{\max} = A + A_{\text{range}}$ |
| $\text{coss}(a\vert x_1, y_1, x_2, y_2, x_3, y_3)$ | Cosine-smoothing function | For a given age $a$ (the midpoint age of age group $i$) the function evaluates to $y_1$ for $a \leq x_1$, to $y_2$ for $a = x_2$, and to $y_3$ for $a \geq x_3$. Values of $a$ between $x_1$ and $x_2$ are interpolated between $y_1$ and $y_2$, and values of $a$ between $x_2$ and $x_3$ are interpolated between $y_2$ and $y_3$, where the interpolation takes the shape of a cosine curve between $-\pi$ and $\pi$. |

**Supplementary Table 2.** Details on mixing matrices used in the study.

| Location | Mixing matrix details |
| --- | --- |
| Wuhan City, China | We used mixing matrices measured in Shanghai in 2017/2018<sup>6</sup>, adapted to the demographics of Wuhan prefecture. This implicitly assumes that Shanghai mixing patterns are representative of large cities in China. |
| Regions of China: Anhui, Guangdong, Guangxi, Hubei, Hunan, Jiangsu, Jiangxi, Jilin Shaanxi, Shandong, Sichuan, Tianjin, Zheijiang provinces; Beijing, Shanghai. | We used mixing matrices measured in Shanghai in 2017/2018<sup>6</sup>, adapted to the demographics of each province / city. |
| Regions of Italy: Lombardia, Piemonte, Trento Veneto, Friulli Venezia Giulia, Liguria, Emilia-Romagna, Toscana, Marche, Lazio, Campania, Puglia regions; Milan. | We used mixing matrices measured in Italy in 2005/2006<sup>7</sup>, adapted to the demographics of each region / city. This assumes that these contact patterns will still be representative of contact patterns in 2020. |
| Ontario, Canada | We used synthetic contact matrices, generated based on demographic information about the country<sup>8</sup>. |
| Japan | We used synthetic contact matrices, generated based on demographic information about the country<sup>8</sup>. |
| Singapore | We used synthetic contact matrices based on demographic information about the country<sup>8</sup>. |
| South Korea | We used synthetic contact matrices based on demographic information about the country<sup>8</sup>. |
| Birmingham, UK | We used mixing matrices measured in the UK in 2005/2006<sup>7</sup>, adapted to the demographics of Birmingham. This assumes that these contact patterns will still be representative of contact patterns in 2020. |
| Bulawayo, Zimbabwe | We used mixing matrices measured in Manicaland, Zimbabwe in 2013<sup>9</sup>, adapted to the demographics of Bulawayo. This implicitly assumes that Manicaland mixing patterns are representative of Bulawayo. |
| 150 capital cities | We used synthetic contact matrices, generated based on demographic information about each country<sup>8</sup>. |

**Supplementary Table 3.**
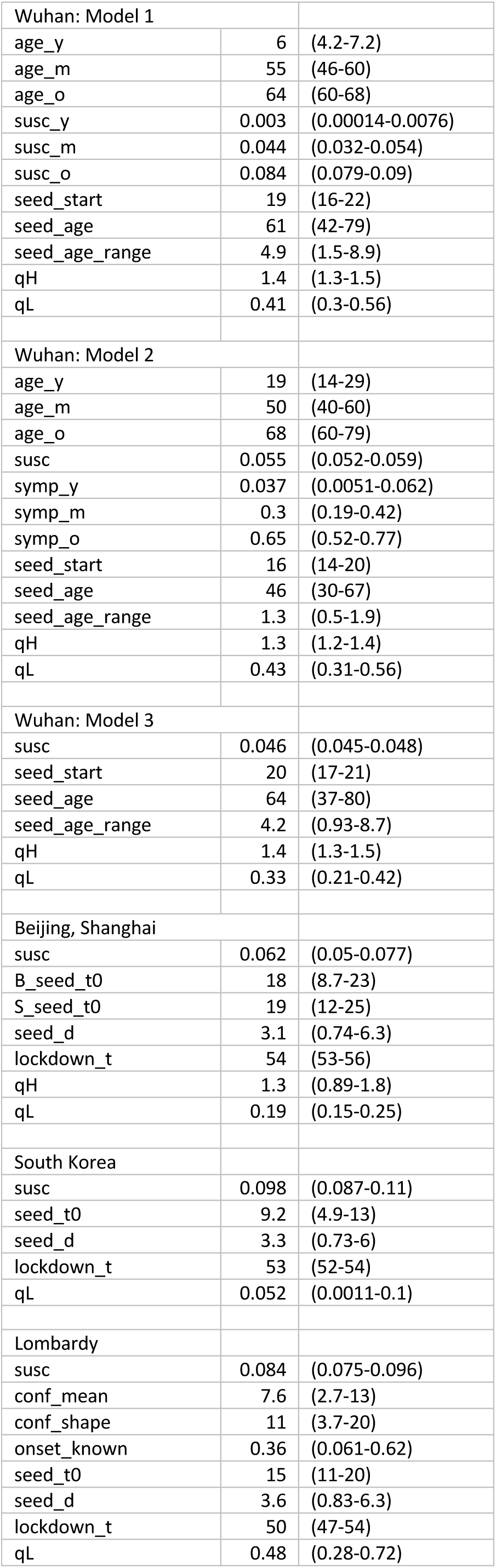
Posterior means and 95% HDIs from fitting the dynamic transmission model (Figs. 1 and 2, main text).

## Data Availability

All data used for the analysis are publicly available. Analysis code will be made available on Github.

## Notes

### Competing Interest Statement

The authors have declared no competing interest.

### Funding Statement

We acknowledge the following for funding: NGD: National Institutes of Health Research (HPRU-2012-10096). PK, YL, KP, MJ: This research was partly funded by the Bill & Melinda Gates Foundation (INV-003174). YL, MJ: This research was partly funded by the National Institute for Health Research (NIHR) (16/137/109) using UK aid from the UK Government to support global health research. The views expressed in this publication are those of the author(s) and not necessarily those of the NIHR or the UK Department of Health and Social Care. RME: HDR UK (grant: MR/S003975/1).
The members of the CMMID COVID-19 working group and the funding they acknowledge are: Carl A B Pearson, Billy J Quilty (NIHR 16/137/109), Adam J Kucharski (Wellcome Trust grant: 206250/Z/17/Z), Hamish Gibbs (funded by the Department of Health and Social Care using UK Aid funding and is managed by the NIHR. The views expressed in this publication are those of the author(s) and not necessarily those of the Department of Health and SocialCare (ITCRZ 03010), Samuel Clifford (Wellcome Trust grant: 208812/Z/17/Z), Amy Gimma (Global Challenges Research Fund (GCRF) for the project "RECAP" managed through RCUK and ESRC (ES/P010873/1), Kevin van Zandvoort (supported by Elrha’s Research for Health in Humanitarian Crises (R2HC) Programme, which aims to improve health outcomes by strengthening the evidence base for public health interventions in humanitarian crises. The R2HC programme is funded by the UK Government (DFID), the Wellcome Trust, and the UK National Institute for Health Research (NIHR), James D Munday (Wellcome Trust grant: 210758/Z/18/Z), Charlie Diamond (NIHR 16/137/109), W John Edmunds, Joel Hellewell (Wellcome Trust grant: 210758/Z/18/Z), Timothy W Russel (Wellcome Trust grant: 206250/Z/17/Z), Sam Abbott (Wellcome Trust grant: 210758/Z/18/Z), Sebastian Funk (Wellcome Trust grant: 210758/Z/18/Z), Nikos I Bosse, Fiona Sun (NIHR EPIC grant 16/137/109), Stefan Flasche (Wellcome Trust grant: 208812/Z/17/Z), Alicia Rosello (NIHR grant: PR-OD-1017-20002), Christopher I Jarvis (Global Challenges Research Fund (GCRF) project ‘RECAP’ managed through RCUK and ESRC (ES/P010873/1)), RMGJH (European Research Commission Starting Grant: #757699).

